# Improvement and Multi-Population Generalizability of a Deep Learning-Based Chest Radiograph Severity Score for COVID-19

**DOI:** 10.1101/2020.09.15.20195453

**Authors:** Matthew D. Li, Nishanth T. Arun, Mehak Aggarwal, Sharut Gupta, Praveer Singh, Brent P. Little, Dexter P. Mendoza, Gustavo C.A. Corradi, Marcelo S. Takahashi, Suely F. Ferraciolli, Marc D. Succi, Min Lang, Bernardo C. Bizzo, Ittai Dayan, Felipe C. Kitamura, Jayashree Kalpathy-Cramer

**Affiliations:** Athinoula A. Martinos Center for Biomedical Imaging, Department of Radiology, Massachusetts General Hospital, Harvard Medical School, Boston, MA, USA; Division of Thoracic Imaging and Intervention, Department of Radiology, Massachusetts General Hospital, Harvard Medical School, Boston, MA, USA; Diagnósticos da América SA (DASA), São Paulo, Brazil; Department of Diagnostic Imaging, Universidade Federal de São Paulo, São Paulo, Brazil; Division of Emergency Radiology, Department of Radiology, Massachusetts General Hospital, Harvard Medical School, Boston, MA, USA; MGH and BWH Center for Clinical Data Science, Mass General Brigham, Boston, MA, USA

**Keywords:** COVID-19, chest radiograph, deep learning, artificial intelligence, generalizability

## Abstract

**Purpose:** To improve and test the generalizability of a deep learning-based model for assessment of COVID-19 lung disease severity on chest radiographs (CXRs) from different patient populations.

**Materials and Methods:** A published convolutional Siamese neural network-based model previously trained on hospitalized patients with COVID-19 was tuned using 250 outpatient CXRs. This model produces a quantitative measure of COVID-19 lung disease severity (pulmonary x-ray severity (PXS) score). The model was evaluated on CXRs from four test sets, including 3 from the United States (patients hospitalized at an academic medical center (N=154), patients hospitalized at a community hospital (N=113), and outpatients (N=108)) and 1 from Brazil (patients at an academic medical center emergency department (N=303)). Radiologists from both countries independently assigned reference standard CXR severity scores, which were correlated with the PXS scores as a measure of model performance (Pearson r). The Uniform Manifold Approximation and Projection (UMAP) technique was used to visualize the neural network results.

**Results:** Tuning the deep learning model with outpatient data improved model performance in two United States hospitalized patient datasets (r=0.88 and r=0.90, compared to baseline r=0.86). Model performance was similar, though slightly lower, when tested on the United States outpatient and Brazil emergency department datasets (r=0.86 and r=0.85, respectively). UMAP showed that the model learned disease severity information that generalized across test sets.

**Conclusions:** Performance of a deep learning-based model that extracts a COVID-19 severity score on CXRs improved using training data from a different patient cohort (outpatient versus hospitalized) and generalized across multiple populations.

## INTRODUCTION

Chest radiographs (CXRs) are routinely obtained in symptomatic patients with suspected or confirmed coronavirus disease 2019 (COVID-19) infection. While CXRs have limited sensitivity for the diagnosis of COVID-19,^1–3^ the severity of radiographic lung findings has been associated with worse clinical outcomes.^4–6^ Deep learning-based techniques have been used to automate the extraction of measures of lung disease severity from CXR image data, which correlate with manual scores of disease severity by radiologists and can be potentially used for patient risk stratification.^7–12^ These techniques are promising; however, the performance of CXR deep learning models are known to show variable generalization on external data.^13^ Thus, validation on data from different sources and patient populations is essential before such models can be deployed in clinical practice.

In this study, we aimed to improve and test the generalizability of a previously published deep learning-based model for automated assessment of COVID-19 pulmonary disease severity, the Pulmonary X-Ray Severity (PXS) score model.^7^ A limitation of the original model was that it was trained and tested on CXRs from patients hospitalized with COVID-19, who tend to have more severe disease compared to the general population infected by COVID-19. In addition, portable anterior-posterior (AP) CXRs are overrepresented compared to standard posterior-anterior (PA) CXRs, which may be more common in outpatient settings. In this work, we tuned the PXS score model by training with primarily outpatient CXRs and assessed model performance in comparison to manual radiologist annotations for lung disease severity in four different test sets with different technical and patient characteristics, including CXRs acquired from patients in two countries (United States and Brazil).

## MATERIALS AND METHODS

This retrospective study was reviewed and exempted by the Institutional Review Board of Massachusetts General Brigham (Boston, USA), with waiver of informed consent. The parts of the study involving data from Hospital Santa Paula was approved by the Institutional Review Board of the Universidade Federal de São Paulo (São Paulo, Brazil). The hospitals involved in this study include Massachusetts General Hospital (Hospital 1) (Boston, USA), Hospital Santa Paula(Hospital 2) (São Paulo, Brazil), and Newton Wellesley Hospital (Hospital 3) (Newton, USA), Hospitals 1 and 2 are large academic medical centers, while Hospital 3 is a community hospital in the Boston metropolitan area.

### PXS Score Base Model

For the base model for this study, we used a previously published convolutional Siamese neural network-based model that can extract a continuous measure of lung disease severity from CXRs in patients with COVID-19, the PXS score model.^7^ In brief, a Siamese neural network is composed of twinned subnetworks with identical weights; paired images can be passed as inputs, each image passing to a subnetwork.^14^ The Euclidean distance between the last fully connected layers of the subnetworks can serve as a continuous measure of disease severity similarity between the two input images.^15^ In the original PXS score model, a Siamese neural network composed of twinned DenseNet121 networks^16^ was pre-trained using ∼160,000 anterior-posterior (AP) chest radiographs from the publicly available CheXpert dataset.^17^ The model was then trained using 314 admission CXRs from hospitalized patients with COVID-19 at Hospital 1 annotated by radiologists using a manual scoring system for lung disease severity.^7^ During model inference, the image-of-interest was compared to a pool of normal CXRs from CheXpert, and the median of the Euclidean distances between the image-of-interest and each normal CXR served as the PXS score. Please refer to the previously published work for the technical details of this implementation.^7^ See Figure 1 for a study design schematic.

**Figure 1.**
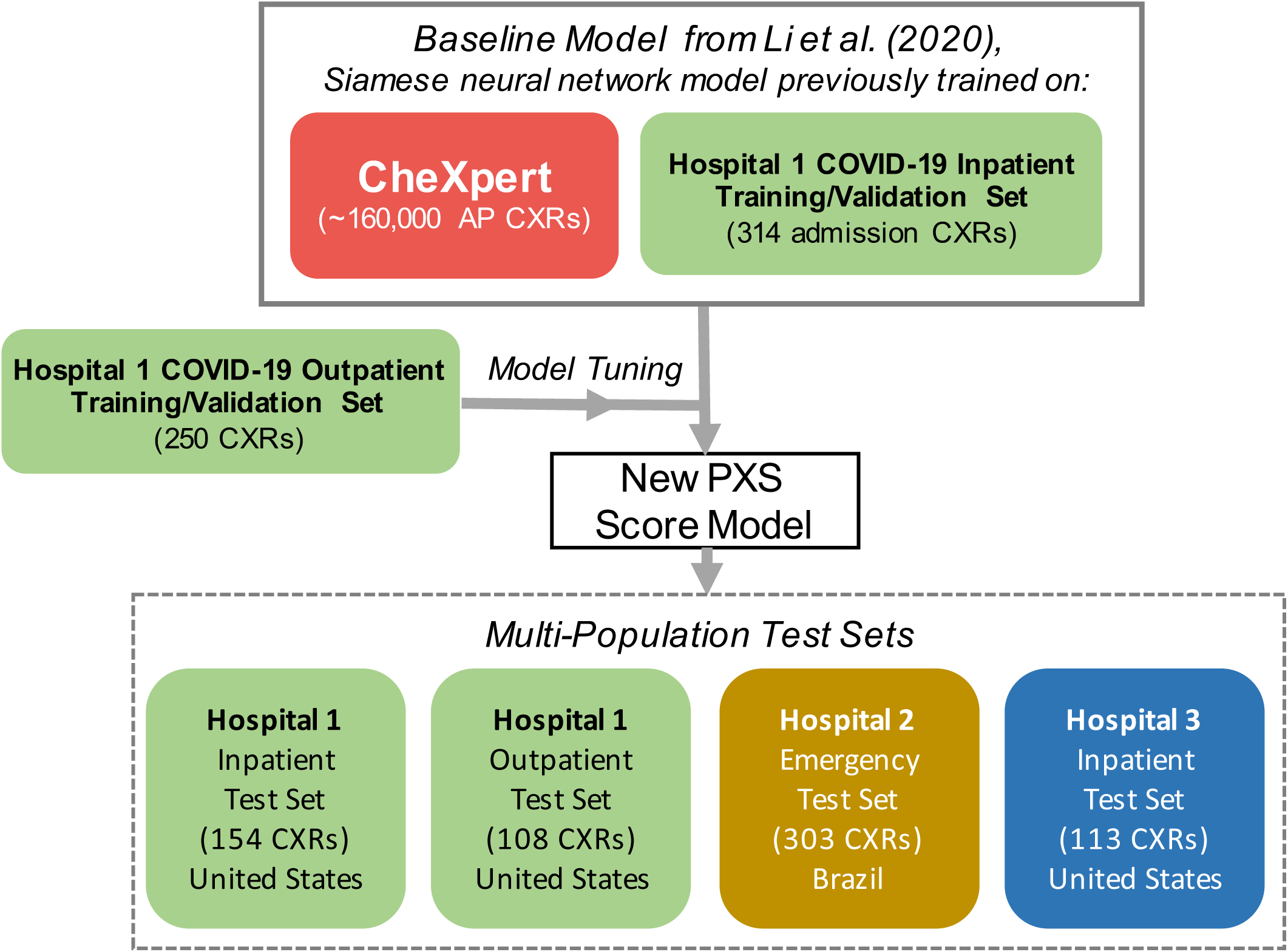
Schematic of study design. Previously published Siamese neural network-based model for extracting lung disease severity from CXRs^7^ was tuned using new CXR data and evaluated in four test sets.

### Chest Radiograph Data

We assembled two new CXR DICOM datasets for this study:

#### (1) Hospital 1 Outpatient Dataset

This dataset was composed of 358 CXRs from 349 unique patients who presented for outpatient imaging at urgent care or respiratory illness clinics associated with Hospital 1 and tested positive for COVID-19 by nasopharyngeal swab RT-PCR obtained at their outpatient visit from March 15, 2020 to April 15, 2020. Raw DICOM data for the frontal view CXRs was extracted and anonymized directly from the institutional PACS. This dataset was composed of mostly CXRs acquired in the posterior-anterior (PA) position (342, 96%), with the remainder acquired in the anterior-posterior (AP) position (16, 4%). Some radiographs in this data set overlapped with the original PXS score model training data set (22, 6%) because the outpatient CXR was used as the admission CXR in some patients.^7^ Thus, in partitioning this outpatient CXR dataset, we included the overlapping radiographs in the planned training/validation partition and then randomly allocated the remaining CXRs up to a 70:30 distribution (250 for training/validation and 108 for testing). The training/validation partition was then randomly partitioned 90:10 (225 for training, 25 for validation). Associated age and sex data were extracted from the electronic health record.

#### (2) Hospital 2 Emergency Test Set

This dataset was composed of 303 CXRs from 242 unique patients who presented to the emergency department with suspected COVID-19 at Hospital 2. These CXRs were sampled from patients from February 1, 2020 to May 30, 2020 with at least one COVID-19 RT-PCR result within ±3 days of the CXR. Sampling was stratified on RT-PCR test results, so that 70% of CXRs in the dataset would have at least one positive associated test and 30% would have all negative tests. In addition, a subset of patients was permitted to have multiple CXRs in the dataset (49 with 2 CXRs, 6 with 3 CXRs). Raw DICOM data for the frontal view CXRs was extracted and anonymized from the institutional PACS. The AP versus PA view position was not available in the DICOM metadata for this data. Age and sex data were extracted from the electronic health record.

In addition to these two data sets that were created for this study, we also used previously published data sets for model testing, including 154 admission CXRs from 154 unique patients hospitalized for COVID-19 at Hospital 1 (**Hospital 1 Inpatient Test Set**) and 113 admission CXRs from 113 unique patients hospitalized for COVID-19 at Hospital 3 (**Hospital 3 Inpatient Test Set**).^7^ X-ray equipment manufacturer information from all four test sets were extracted from the DICOM metadata tags.

Raw pixel data from the CXR DICOMs used in training, validation, and testing were pre-processed using the same steps as used in the baseline PXS score model,^7^ including conversion to 8-bit, correction of photometric inversion, histogram equalization, and conversion to a JPEG file.

### Radiologist Annotations for Lung Disease Severity

We used a manual scoring system for COVID-19 lung disease severity on CXRs previously used for training of the PXS score model, which is a modified version of the Radiographic Assessment of Lung Edema scoring system (mRALE).^7,18^ In brief, from the frontal view of the CXR, each lung is assigned a score from 0 to 4 for extent of consolidation or ground glass/hazy opacities (up to 0, 25, 50, 75, 100%) and a score from 1 to 3 for overall density (hazy, moderate, dense). The sum of the products of the extent and density scores for each lung is the mRALE score (range from 0-24). Higher mRALE scores have been associated with worse clinical outcomes in COVID-19.^5^ Two diagnostic radiologists with thoracic subspecialty expertise (B.P.L., D.P.M.) from Hospital 1 independently annotated the 358 CXRs from the Hospital 1 Outpatient Dataset for mRALE, viewing the images on a diagnostic PACS viewer. Three diagnostic radiologists with non-thoracic subspecialty training (G.C.A.C., M.S.T., S.F.F.) from Hospital 2 independently annotated the 303 CXRs from the Hospital 2 Emergency Test Set for mRALE, viewing the images using the MD.ai annotation platform (New York, United States). The average of the rater mRALE scores served as the mRALE score for each CXR. All raters had previously rated 10 CXRs using mRALE with feedback on their scores, though the Hospital 1 raters had more experience, previously rating ∼300 studies independently.

To assess the correlation between the radiologists from both hospitals in applying the mRALE score, the two thoracic radiologists from Hospital 1 rated a subset of 69 studies from the Hospital 2 dataset in PACS viewers. This subset was composed of studies with mRALE ≥ 3.0 assigned by the Hospital 3 raters, in order to focus re-assessment on abnormal lungs, rather than normal / near-normal lungs.

### PXS Score Model Re-Training

The base PXS score Siamese neural network model was re-trained (“tuned”) using the 250 CXR training/validation partition of the Hospital 1 Outpatient Dataset, using the same training strategy with mean square error (MSE) loss as previously reported.^7^ In brief, random CXR image pairs were fed to the Siamese neural network. The difference between the Euclidean distance between the final fully connected layers of the network and the absolute difference in mRALE scores between the two input images served as the “error” for the MSE loss function. During model training and validation, 1600 and 200 input image pairs were randomly sampled per epoch, respectively. For training, input images were randomly rotated ± 5° and then randomly cropped to a scale of 0.8-1 and resized to 320 x 320 pixels.

For validation, input images were resized to 336 ⨯ 336 pixels and center cropped to 320 ⨯ 320 pixels. The model training was implemented in Python (version 3.6.9) with the Pytorch package (version 1.5.0), using the Adam optimizer^19^ (initial learning rate = 0.00002, β_1_ = 0.9, β_2_ = 0.999). Training/validation batch sizes of 8 and early stopping at 7 epochs without improvement in validation loss were set. The lowest validation loss model was saved for evaluation. The code used for model training is available at *https://github.com/QTIM-Lab/PXS-score*.

### PXS Score Model Inference

The PXS score for an image-of-interest is the median of Euclidean distances calculated from paired image inputs passed through the Siamese neural network, where each paired image input consists of the image-of-interest and an image from a pool of *N* normal CXRs. In this study, we created a set of 15 manually curated normal chest x-rays with varying body habitus and field-of-view from CXRs from Hospital 1 to serve the pool of normal CXRs (age range 18-72 years, 7 women and 8 men).

In some CXR images, primarily in the Hospital 3 dataset, large black borders may surround the actual CXR. Immediately before the histogram normalization step described in the pre-processing step described above, a Python script for automated rectangular cropping for black borders was applied to the image (i.e. border pixels with normalized values <2 were cropped). Code used for model inference and this cropping step is also available at the GitHub link above.

### Statistics / Data Visualization

To evaluate differences in sex between the datasets, we used the Chi-square test. To evaluate differences in age and mRALE scores (treated as a continuous variable from 0-24), we used the Kruskal-Wallis test and post-hoc Mann-Whitney tests (two-sided). Interrater correlations for mRALE labeling and correlations between PXS score and mRALE were assessed using Pearson correlations (r). Statistical tests were performed using the *scipy* Python package (version 1.1.0), with an *a priori* threshold for statistical significance set at P<0.05.

The *Seaborn* Python package (version 0.10.0) was used for scatterplot data visualizations. To perform dimensionality reduction for visualizing the neural network results, we used the Python implementation of UMAP (Uniform Manifold Approximation and Projection) (version 0.4.2) (number of neighbors = 20, minimum distance = 0.6, metric = correlation).^20,21^ Each test set image was passed through a single subnetwork of the Siamese neural network and the last fully connected layer (1000 nodes in DenseNet121) from each image was used as an input for UMAP.

## RESULTS

### Chest Radiograph Dataset Characteristics

The Hospital 1 Outpatient Dataset (including training/validation and test partitions) and Hospital 2 Emergency Test Set characteristics are summarized in Table 1. The Hospital 1 Inpatient Test Set and Hospital 3 Inpatient Test Set characteristics were previously published.^7^ There were significantly different age distributions between the test sets (p=0.003) (Figure 2A).The Hospital 1 Outpatient Test Set patient ages were significantly lower compared to the Hospital 1 and 3 Inpatient Test Sets (median 53 versus 59 years, p=0.003, and median 53 versus 74 years, p<0.001, respectively). The Hospital 2 Emergency Test Set ages were significantly lower than the Hospital 1 Outpatient Test Set ages (median 41 versus 53 years, p<0.001). There was a significantly higher proportion of CXRs from women in the dataset from Brazil compared to the combined datasets from the United States (58% versus 45%, p=0.001). The X-ray equipment used to obtain these CXRs came from a variety of manufacturers that differed by dataset (Table 2).

**Table 1.** Summary of dataset characteristics and radiologist mRALE scores. N, Number; Q1-Q3, Quartile 1 to Quartile 3 (i.e. interquartile range).

|  | Hospital 1 Outpatient Dataset<br>(UNITED STATES)<br>Patients presenting for outpatient imaging who<br>tested positive by COVID-19 RT-PCR |  |  |  | Hospital 2 Emergency Test Set<br>(BRAZIL)<br>Patients presenting to emergency department<br>with suspected COVID-19 |  |  |  |
| --- | --- | --- | --- | --- | --- | --- | --- | --- |
|  | All | Training/<br>Validation<br>Set | Outpatient<br>Test Set | p-value<sup>a</sup> | All | RT-PCR<br>positive | RT-PCR<br>negative | p-value<sup>b</sup> |
| **CXRs, N** | 358 | 250 | 108 |  | 303 | 203 | 100 |  |
| **Unique Patients, N** | 349 | 248 | 106 |  | 242 | 167 | 75 |  |
| **Age (years),**<br>**median (Q1-Q3)** | 53 (41-64) | 52 (41-65) | 53 (41-63) | 0.9 | 41 (33-52) | 40 (33-50) | 44 (33-52) | 0.2 |
| **Sex, N women (%)** | 186 (52%) | 132 (53%) | 54 (50%) | 0.7 | 175 (58%) | 113 (56%) | 62 (62%) | 0.4 |
| **mRALE, median**<br>**(Q1-Q3)** | 1.0 (0-3.5) | 1.0 (0-3.0) | 1.0 (0-4.5) | 0.2 | 0.3 (0-2.7) | 0.3 (0-2.8) | 0.3 (0-1.8) | 0.6 |
| **mRALE, N (%)** |  |  |  |  |  |  |  |  |
| **mRALE = 0** | 123 (34%) | 88 (35%) | 35 (32%) |  | 122 (40%) | 84 (41%) | 38 (38%) |  |
| **0 < mRALE ≤ 4** | 164 (46%) | 122 (49%) | 42 (39%) |  | 126 (42%) | 78 (38%) | 48 (48%) |  |
| **4 < mRALE ≤ 10** | 58 (16%) | 30 (12%) | 28 (26%) |  | 29 (10%) | 22 (11%) | 7 (7%) |  |
| **mRALE > 10** | 13 (4%) | 10 (4%) | 3 (3%) |  | 26 (9%) | 19 (9%) | 7 (7%) |  |

<sup>a</sup>p-value for comparison of internal test set with training/validation set; <sup>b</sup>p-value for comparison of patients who tested positive versus negative by COVID-19 RT-PCR.

**Table 2.** Summary of x-ray equipment manufacturers extracted from DICOM metadata.

| **Dataset** | **Manufacturer (Headquarters)** | **Number of CXRs** |
| --- | --- | --- |
| Hospital 1 Inpatient Test Set (United States) | Agfa (Mortsel, Belgium)<br>GE Healthcare (Chicago, USA)<br>Varian (Palo Alto, USA)<br>Not available | 136<br>1<br>4<br>13 |
| Hospital 1 Outpatient Test Set (United States) | Agfa (Mortsel, Belgium) | 108 |
| Hospital 2 Emergency Test Set (Brazil) | Fujifilm Corporation (Tokyo, Japan) | 303 |
| Hospital 3 Inpatient Test Set (United States) | Agfa (Mortsel, Belgium)<br>Carestream (Rochester, USA)<br>Kodak (Rochester, USA)<br>Philips (Amsterdam, Netherlands)<br>Siemens (Munich, Germany) | 33<br>72<br>2<br>2<br>2 |

**Figure 2.**
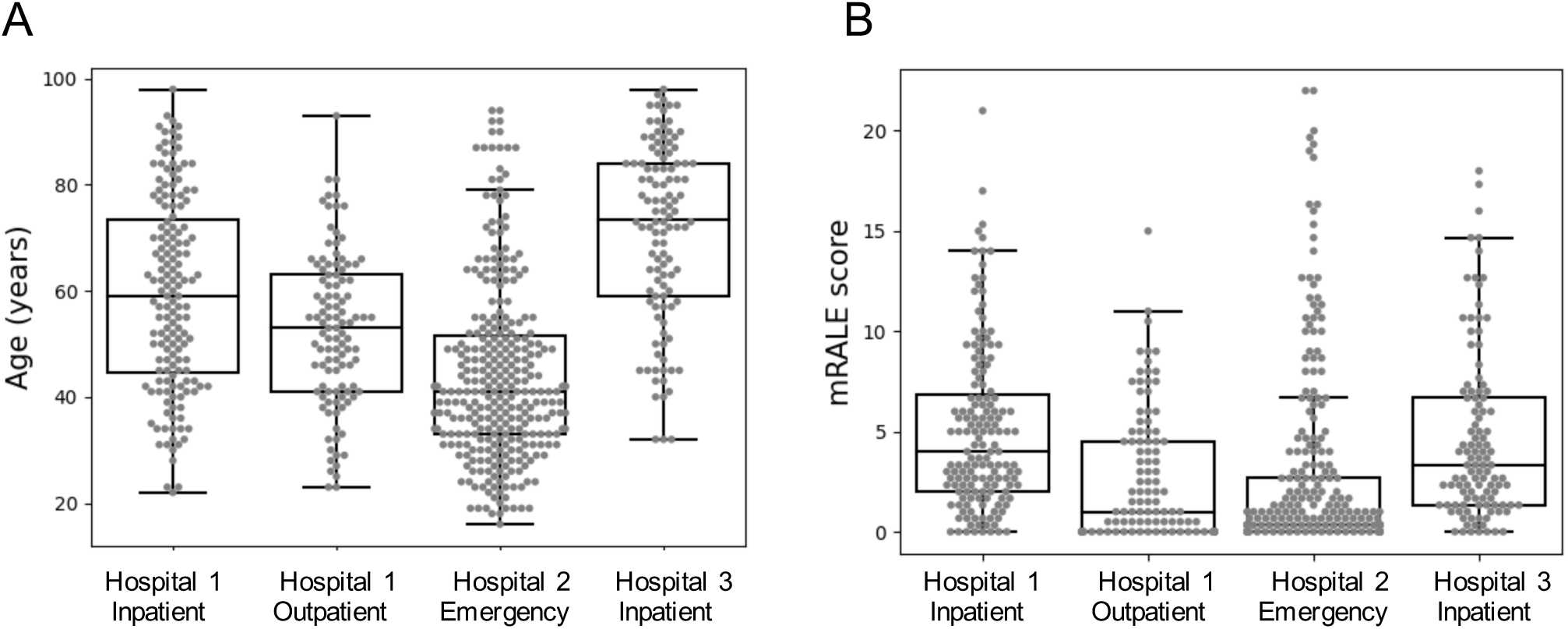
Boxplots show variable distributions in patient age (A) and lung disease severity by mRALE score (B) in the different CXR test sets. Boxplots show the median and interquartile range (IQR), where the whiskers extend up to 1.5 x IQR.

### Radiologist Annotations of COVID-19 Lung Disease Severity

The correlation between the two raters at Hospital 1 for assigning mRALE scores to the 358-CXR Hospital 1 Outpatient Dataset was high (r=0.89, p<0.001). The correlation between the three raters at Hospital 2 for assigning mRALE scores to the 303-CXR Hospital 2 Emergency Test Set was lower (r=0.85, 0.81, and 0.84, for each pairwise comparison; p<0.001 in all comparisons). In the 69-CXR subset of the Hospital 2 Emergency Test Set that the Hospital 1 raters also evaluated, the correlation between the average Hospital 1 and average Hospital 2 rater mRALE scores was 0.86 (p<0.001). However, the individual Hospital 2 rater mRALE scores showed variable correlation with the average Hospital 1 raters (r=0.65, 0.75, 0.86).

There were significantly different mRALE distributions between the test sets (p=0.011) (Figure 2B). The mRALE scores were significantly lower in the Hospital 1 Outpatient Dataset compared to the Hospital 1 and 3 Inpatient Test Sets (median 1.0 versus 4.0, p<0.001, and median 1.0 versus 3.3, p<0.001). The mRALE scores in the Hospital 2 Emergency Test Set were significantly lower compared to each of the other test sets (all p<0.001).

### Improved Deep Learning Model Performance and Testing of Generalizability

The PXS score model tuned using the Hospital 1 Outpatient Training/Validation Set showed improvements in correlation between the model output (PXS score) and radiologist-determined mRALE scores in the Hospital 1 Inpatient and Hospital 3 Inpatient Test Sets (r=0.88 and r=0.90, respectively, p<0.001; increased from r=0.86 and r=0.86 using the baseline model) (Figures 3A and 3D)

**Figure 3.**
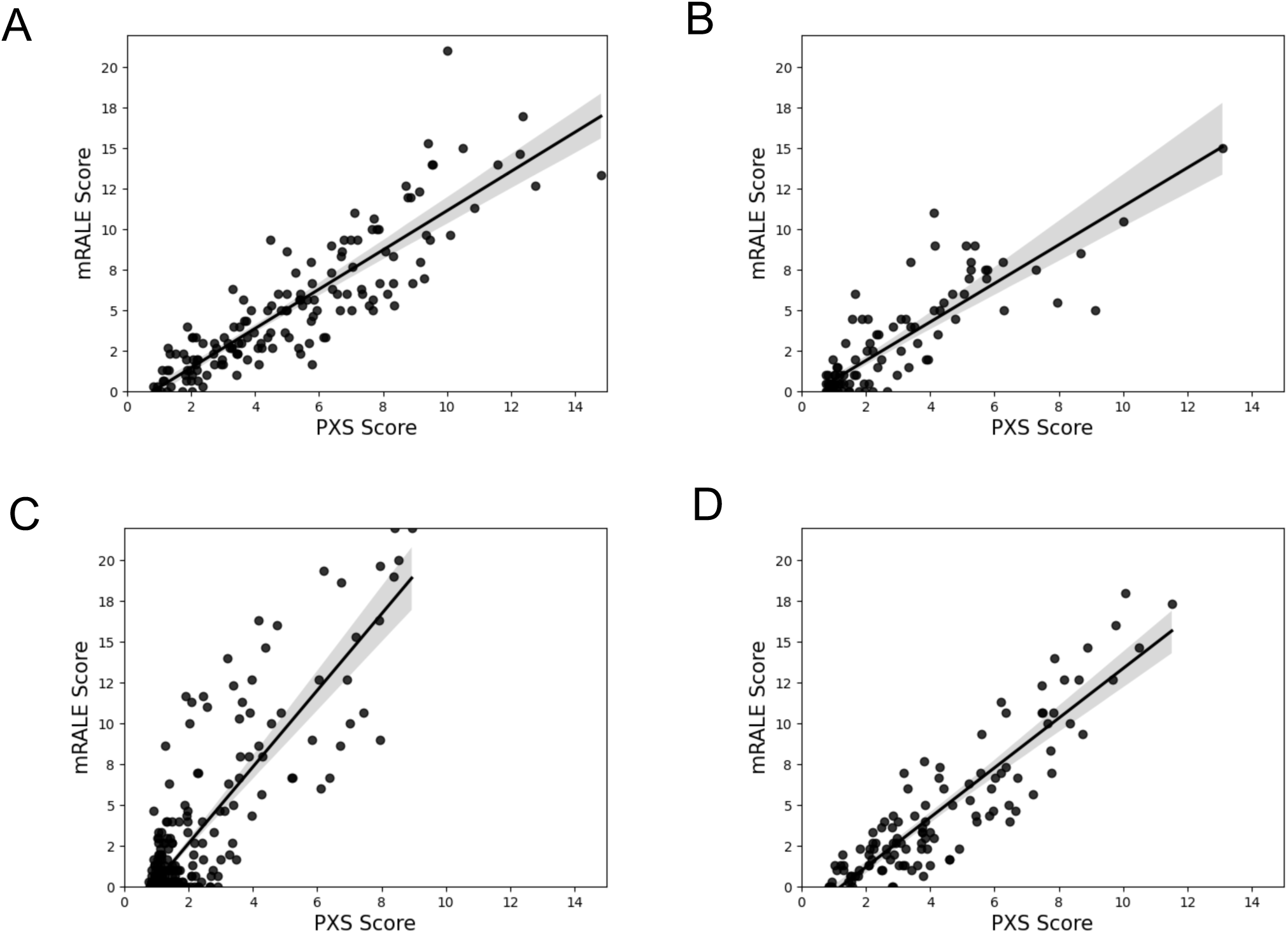
Scatterplots show the correlation between radiologist-determined mRALE score and the deep learning-based PXS score in the Hospital 1 Inpatient Test Set (r=0.88) (A), Hospital 1 Outpatient Test Set (r=0.86) (B), Hospital 2 Emergency Test Set (r=0.85) (C), and Hospital 3 Inpatient Test Set (r=0.90) (A). Linear regression 95% confidence intervals are shown in each scatterplot.

We further tested this tuned PXS score model on the Hospital 1 Outpatient and Hospital 2 Emergency Test Sets, which showed that the model could generalize to these additional datasets (r=0.86 and r=0.85 respectively, p<0.001) (Figures 3B and 3C). However, there was a steeper slope for the regression on the Hospital 2 Emergency Test Set data (slope=2.3) versus the slope of the other test sets (in aggregate, slope=0.6). While the model learned a measure of disease severity as evidenced by the significant correlation between mRALE and PXS scores, for this specific test set, the relationship between mRALE and PXS was scaled differently.

### Visualizing Test Set Relationships Using Dimensionality Reduction

When CXRs from all four test sets were analyzed in aggregate (total N=678), UMAP showed that the CXRs appear to cluster principally in relation to similar disease severity (PXS and mRALE scores) (Figures 4A and 4B). Contrastingly, there was substantial overlap between the CXRs from different test sets (Figure 4C). These findings support the finding that the PXS score model learned a generalizable representation of lung disease severity. However, the normal or near-normal severity CXRs appear to have a larger cluster in the Hospital 2 Emergency Test Set compared to the other test sets (Figure 4C). We visually inspected these images and did not find a systematic perceptible difference in view position, body habitus, heart size or x-ray exposure.

**Figure 4.**
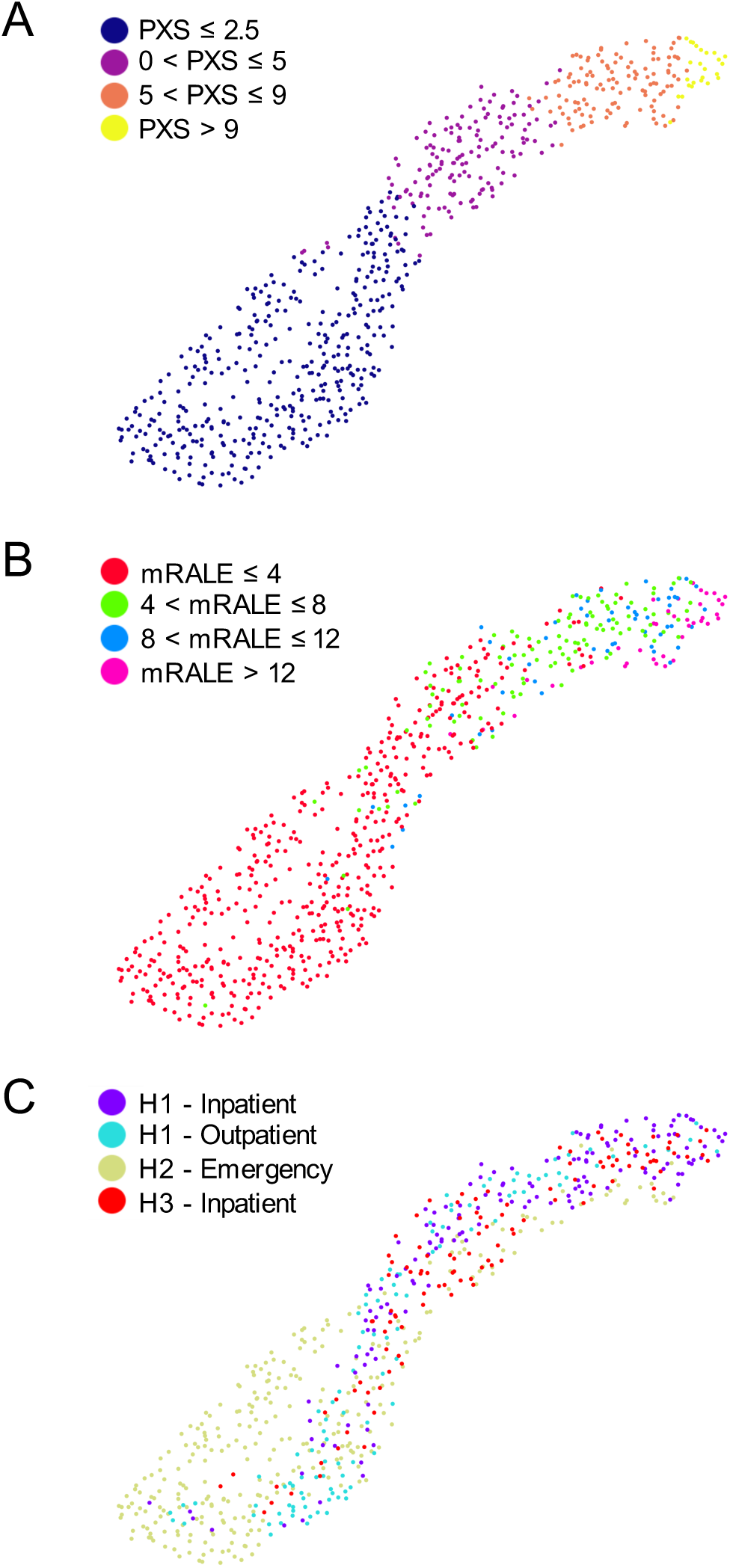
Dimensionality reduction using UMAP shows the relationships between CXR data passed through the deep learning-based PXS score model from all four test sets (total N=678), color coded for PXS score (A), mRALE score (B), and test set (C). For the legend in (C), *H* indicates Hospital. Across the different test sets, a representation of lung disease severity is learned by the PXS score model.

## DISCUSSION

We improved the performance of the deep learning-based PXS score model for assessment of a quantitative measure of COVID-19 lung disease severity on CXRs and tested its generalizability on four test sets reflective of different populations from the United States and Brazil. The PXS model was originally trained using admission CXRs from hospitalized COVID-19 patients.^7^ In this study, we found that tuning the deep learning model using CXR data from outpatients improved model performance on the test sets from hospitalized patients. This may be because the model was able to learn from a different distribution of data, as outpatients have typically less severe lung disease and also different view positions compared to admitted patients. Based on the correlation of the model results with manual radiologist annotations for lung disease severity in multiple test sets, the PXS score model does appear to generalize to different patient cohorts. Further supporting this conclusion, a dimensionality reduction technique showed that the CXRs from different test sets cluster primarily by lung disease severity, as opposed to by test set source.

While the correlation between the radiologist-determined severity score (mRALE) and the deep learning-based PXS score was generalizable between test sets, there was a difference in the mRALE/PXS slope between the test sets from the United States and Brazil. Thus, differences in calibration between mRALE and PXS scores may occur for CXRs coming from different sources and this needs to be considered before the use of such a model clinically. This phenomenon could be due to systematic differences in x-ray equipment manufacturers and acquisition technique (including parameters like x-ray tube voltage and current), which can alter the properties of tissue contrast in the image. Subjectively, our radiologist raters found a perceptible difference in exposure/contrast in the images from Brazil versus the United States. The PXS score model attempts to address this issue using histogram normalization, but this transformation may not eliminate all systematic differences. Model training on data from an increased variety of vendors could help address this calibration issue and is a direction of future research.

In spite of the calibration issue, the finding that the PXS score model was able to correlate with manual radiologist annotations at multiple test sites has potential clinical application for the reproducible assessment of COVID-19 lung disease severity at different sites. This reproducible assessment is important because CXR findings have been associated with worse clinical outcomes in patients with COVID-19,^4–6^ which may be useful for clinical risk stratification, and there is interrater variation between radiologist assessments (which will be more pronounced in the “real world” where radiologists are not uniformly trained on the use of a scoring system). Another possible application is for radiologist worklist prioritization, which could help the expedite identification of the sickest patients.^22^

Previous work on developing deep-learning based models to assess COVID-19 lung disease severity on CXRs have shown correlations between various systems of manual radiologist assessments and deep learning outputs, though often without external testing. For example, Cohen et al. split their 94 posterior-anterior CXR dataset 50:50 for training and testing,^12^ Zhu et al. split their 131 portable CXR dataset 80:20 for training and testing,^8^ and Blain et al. reported performance on a 65 CXR dataset using 5-fold cross validation.^10^ On the other hand, work from Amer et al. and Signoroni et al.^9,11^ does include external testing of their deep learning models on CXRs from the Cohen et al. dataset,^12^ and Barbosa et al. also perform external testing on an 86 CXR dataset.^23^ Future work in this field should continue to include assessment of model performance across multiple sites, to characterize generalizability for different x-ray acquisition techniques and patient populations before these artificial intelligence-based tools can be deployed for possible clinical use.

There are limitations to this study. First, the reference standard label used for disease severity assessment on CXRs is determined by radiologists, which has inherent variability. We used the average of multiple radiologist raters for the reference standard to decrease the variability in this study. However, other reference standards such as CT-derived scores may be promising, as has been found using digitally reconstructed radiographs from CT.^23^ Second, while studying the technical properties of deep learning-based models like PXS score is necessary, making such CXR-based severity scores clinically useful in addressing the COVID-19 pandemic is a different avenue of important research. Future work into how radiologists and other clinicians can use the PXS score (and other developed lung disease severity scores) to guide patient management or workflows will be essential to deliver value.

## Data Availability

The data used in this study contained protected patient health information and are only available under the respective IRBs from Mass General Brigham and Universidade Federal de Sao Paulo.

## ACKNOWLEDGMENTS

We thank members of the QTIM Lab at Massachusetts General Hospital and the MGH and BWH Center for Clinical Data Science for their support in this work.

